# CSF metabolites associated with biomarkers of Alzheimer’s disease pathology

**DOI:** 10.1101/2022.07.20.22277523

**Authors:** Ruocheng Dong, Qiongshi Lu, Hyunseung Kang, Ivonne Suridjan, Gwendlyn Kollmorgen, Norbert Wild, Yuetiva Deming, Carol A. Van Hulle, Rozalyn M. Anderson, Henrik Zetterberg, Kaj Blennow, Cynthia M. Carlsson, Sanjay Asthana, Sterling C. Johnson, Corinne D. Engelman

## Abstract

**INTRODUCTION:** Metabolomics technology facilitates studying associations between small molecules and disease processes. Correlating metabolites in cerebrospinal fluid (CSF) with Alzheimer’s disease (AD) CSF biomarkers may elucidate additional changes that are associated with early AD pathology and enhance our knowledge of the disease.

**METHODS:** The relative abundance of untargeted metabolites was assessed in 161 individuals. A metabolome-wide association study (MWAS) was conducted between 269 CSF metabolites and protein biomarkers reflecting brain amyloidosis, tau pathology, neuronal and synaptic degeneration, and astrocyte or microglial activation and neuroinflammation. Linear mixed-effects regression analyses were performed with random intercepts for sample relatedness and repeated measurements and fixed effects for age, sex, and years of education. The metabolome-wide significance was determined by a false discovery rate threshold of 0.05. The significant metabolites were replicated in 154 independent individuals. Mendelian randomization was performed using genome-wide significant single nucleotide polymorphisms from a CSF metabolites genome-wide association study.

**RESULTS:** MWAS results showed several significantly associated metabolites for all the biomarkers except Aβ42/40 and IL-6. Genetic variants associated with metabolites and Mendelian randomization analysis provided evidence for a causal association of metabolites for soluble triggering receptor expressed on myeloid cells 2 (sTREM2), amyloid β (Aβ40), α-synuclein, total tau, phosphorylated tau, and neurogranin, for example, palmitoyl sphingomyelin (d18:1/16:0) for sTREM2, and erythritol for Aβ40 and α-synuclein.

**DISCUSSION:** This study provides evidence that CSF metabolites are associated with AD-related pathology, and many of these associations may be causal.

## 1. Introduction

The neuropathological changes of Alzheimer’s disease (AD) consist of extracellular amyloid-β (Aβ) plaques and intracellular neurofibrillary tangles of hyperphosphorylated tau proteins in the brain[1]. Well-established core biomarkers that reflect AD pathology and show promising performance in evaluating AD risk and diagnosing AD are the 42 amino acid form Aβ (Aβ42), the ratio of Aβ42/40, phosphorylated tau (P-tau), and total tau (T-tau) in the cerebrospinal fluid (CSF)[2]. However, it has been suggested that other pathophysiology such as neuroinflammation through glial activation and neuronal and synaptic degeneration also contribute to symptomatic AD, and CSF biomarkers of these may provide valuable information about disease progression[2]. Thus, the NeuroToolKit (NTK), a panel of automated CSF immunoassays, was introduced to complement the established core AD biomarkers[3]. The NTK panel includes S100 calcium-binding protein B (S100b), chitinase-3-like protein 1 (YKL-40), and glial fibrillary acidic protein (GFAP) as markers of astrocyte activation; soluble triggering receptor expressed on myeloid cells 2 (sTREM2) and interleukin-6 (IL-6) as markers of microglial activation and inflammation; and neurofilament light (NfL), neurogranin, and α-synuclein as markers of axonal injury and synaptic dysfunction[4].

Untargeted metabolomics technology is a promising approach that can simultaneously identify and quantify a large number of small molecules (<1500 Da, *e.g.*, lipids) in a biological sample[5]. Previous research has shown that metabolomic changes in the human brain and CSF were associated with AD status and AD pathological alterations[6]. For example, Koal *et al.*[6] identified eight metabolites that were significantly increased in the CSF samples with AD-like pathology including an acylcarnitine (C3), two sphingomyelins [SM (d18:1/18:0) and SM (d18:1/18:1)], and five glycerophospholipids (PC aa C32:0, PC aa C34:1, PC aa C36:1, PC aa C38:4, and PC aa C38:6). However, no studies have examined associations between the untargeted CSF metabolome and a broad panel of biomarkers such as the NTK panel. Thus, our study aims to link CSF metabolites with established and developing AD biomarkers with the goals of (1) identifying individual CSF metabolites that are associated with the CSF NTK biomarkers and (2) conducting Mendelian randomization (MR) to determine if the CSF metabolites significantly associated with NTK biomarkers are likely to be in the causal pathway instead of simply changing with, or as a result of, AD biomarker changes.

## 2. Methods

### 2.1 Participants

The Wisconsin Registry for Alzheimer’s Prevention (WRAP) began recruitment in 2001 as a prospective cohort study, with initial follow-up four years after baseline and subsequent ongoing follow-up every two years. WRAP is comprised of initially cognitively-unimpaired, asymptomatic, middle-aged (between 40 and 65) adults enriched for parental history of clinical AD[7]. At each visit, the participants undergo comprehensive medical and cognitive evaluations. Additional details of the study design and methods of WRAP have been described previously[7]. From the WRAP cohort, we identified 161 self-reported non-Hispanic white individuals with longitudinal CSF biomarker and metabolomic data. The sample size for other racial/ethnic groups was too small (n<10) to include in the analyses.

The Wisconsin Alzheimer’s Disease Research Center’s (ADRC) clinical core cohorts started in 2009 and are comprised of well-characterized participants who undergo cognitive testing and physical exams every two years[8]. The Wisconsin ADRC has a cohort of initially cognitively-unimpaired, asymptomatic middle-aged (between 45 and 65) adults with a similar study design to WRAP (the **I**nvestigating **M**emory in **P**reclinical **A**D-**C**auses and Treatments [IMPACT] cohort)[9–11]. From the IMPACT cohort, we identified 154 self-reported non-Hispanic white participants with cross-sectional CSF biomarker and metabolomic data. As with WRAP, the sample size for other racial/ethnic groups was too small (n<10) to include in the analyses.

This study was conducted with the approval of the University of Wisconsin Institutional Review Board, and all participants provided signed informed consent before participation.

### 2.2 CSF sample collection and biomarkers quantification

Fasting CSF samples were collected via lumbar puncture using a Sprotte 25- or 24-gauge spinal needle at the L3/4 or L4/5 interspace with gentle extraction into polypropylene syringes. More details can be found in the previous study[9]. The CSF collection for WRAP and the Wisconsin ADRC followed the same protocol, and the lumbar puncture for both studies was performed by the same group of well-trained individuals.

All CSF samples were batched together and assayed for the NTK biomarkers at the Clinical Neurochemistry Laboratory, University of Gothenburg, using the same lot of reagents, under strict quality control procedures. The immunoassays of Elecsys® Aβ(1-42), P-tau(181P) and T-tau, as well as S100b and IL-6, were performed on a cobas e 601 analyzer[3]. The remaining NTK panel was assayed on a cobas e 411 analyzer including Aβ(1-40), α-synuclein, GFAP, YKL-40, sTREM2, NfL, and neurogranin[3].

### 2.3 CSF metabolomic profiling and quality control

CSF metabolomic analyses and quantification were performed in one batch by Metabolon (Durham, NC) using an untargeted approach, based on Ultrahigh Performance Liquid Chromatography__JTandem Mass Spectrometry platform (UPLC__JMS/MS)[12]. Details of the metabolomic profiling were described in an earlier study[13].

A total of 412 CSF metabolites were identified and quality control procedures were performed. First, 46 metabolites missing for at least 80% of the individuals were excluded. Then the values for each of the remaining metabolites were scaled so that the median equaled 1. Two metabolites with an interquartile range (IQR) of zero were excluded and no metabolites had zero variability between individuals. Log10 transformation was applied to normalize the data. After quality control, 269 metabolites with known biochemical names remained for this investigation. The missing percentage of each metabolite in WRAP and Wisconsin ADRC is available in Supplemental Table 1.

### 2.4 Genotyping and quality control

In the WRAP participants, DNA was extracted from whole blood using the PUREGENE® DNA Isolation Kit, and the concentrations were quantified using the Invitrogen™ Quant-iT™ PicoGreen™ dsDNA Assay Kit. More details can be found in the previous study[13]. Genotyping data were generated by the University of Wisconsin Biotechnology Center using the Illumina Multi-Ethnic Genotyping Array. In the WRAP genetic data, (1) duplicate samples were used to calculate a concordance rate of 99.99%, and discordant genotypes were set to missing; (2) samples missing genotypes for >5% of the single nucleotide polymorphisms (SNPs) were excluded, while SNPs missing in >5% of individuals were also excluded; (3) samples were excluded if the self-reported and genetic sex were inconsistent; (4) SNPs that were not in Hardy-Weinberg equilibrium (HWE; p<3.08E-8) or were monomorphic were removed; (5) individuals that were not of European ancestry were removed due to small sample sizes of other ancestries; (6) the imputation was performed through the Michigan Imputation Server v1.0.328, using the Haplotype Reference Consortium (HRC)[14–16] and the SNPs with a quality score R^2^<0.80, minor allele frequency (MAF)<0.001, or that were out of HWE were excluded; (7) genetic ancestry was assessed by using Principal Components Analysis in Related Samples (PC-AiR) because of the sibling relationships present in the WRAP cohort[13].

Genetic data in the Wisconsin ADRC were generated from DNA extracted from blood samples at baseline and genotyped with either the Infinium OmniExpressExome-8 Kit or the Infinium Global Screening Array-24 Kit. Genetic data for the Wisconsin ADRC underwent the same quality control (QC) and imputation as the WRAP data except samples and SNPs missing in >2% were excluded and HWE threshold was p<1e-6 due to differences in sample sizes and the number of SNPs between the two cohorts.

### 2.5 Statistical analysis

#### 2.5.1 Metabolome-wide association study

A metabolome-wide association study (MWAS) was conducted in the WRAP cohort between 269 individual CSF metabolites and 13 CSF NTK biomarkers using linear mixed-effects regression models with random intercepts to account for repeated measures and family relationships (10 families with two or more siblings) and fixed effects for age at CSF collection, sex, and years of education. Replication of each CSF metabolite significantly associated with one or more biomarkers in WRAP was then conducted in the Wisconsin ADRC cohort using linear regression adjusting for the same covariates. Both Bonferroni and false discovery rate (FDR) methods were used to correct the p-values for multiple testing; the FDR corrected q value was used to determine statistical significance in each analysis. Potential functional pathways of the replicated significant metabolites were identified by pathway analyses using the web-based software Metabo-analyst 5.0[17] based on the Kyoto Encyclopedia of Genes and Genomes (KEGG) Homo sapiens pathway. The hypergeometric test and relative-betweenness centrality were employed to evaluate the pathway importance, and the pathways were considered as important if the impact was ≥0.1.

#### 2.5.2 Prediction performance and elastic net regression

The variance for each biomarker explained by its corresponding significant metabolites was evaluated using r^2^ in the combined cohorts of WRAP and the Wisconsin ADRC. For this analysis, we only included the first available measures of independent participants from WRAP. Since the number of significant metabolites for each biomarker was large and some of the metabolites were highly correlated, elastic net regression[18] was employed to select the important independent metabolites. Then the r^2^ of elastic net-selected metabolites was re-calculated. For each biomarker, we fit three types of models, the (1) base model, which only included the demographics of age, sex, years of education, and cohort, (2) metabolite model, which included the demographics in the base model plus all the replicated significant metabolites, and (3) elastic net-selected metabolite model, which contained the demographics and elastic-net-selected metabolites.

#### 2.5.3 Mendelian randomization

The genome-wide significant SNPs (p < 5 × 10^−8^) from a previous genome-wide meta-analysis of CSF metabolites[19] were extracted for each elastic net-selected metabolite (5863 SNPs for 52 metabolites). These SNPs (or the top 100 SNPs if there were more than 100 genome-wide significant SNPs for a metabolite) were used as instrumental variables (IV) for the metabolite in an MR analysis for each elastic net-selected metabolite-NTK biomarker association pair in the combined WRAP and Wisconsin ADRC cohort. For each MR test, we first checked the strength of the IVs using F statistics. Typically, an IV with an F statistic greater than 10 is considered to be strong, while instruments with F statistics below 10 are considered to be weak[20]. Next, the estimated (or less confounded) beta and p values for the effect of the metabolite on the NTK biomarker were calculated using the two-stage least squares method if the IVs were strong, but using the limited information maximum likelihood (LIML) for IVs that were relatively weak. The confidence intervals (CI) of the point estimates from both LIML and another conditional likelihood ratio (CLR) method, which is robust to weak IVs[21], were compared and only significant results with CIs in the same direction and with a similar range of effect size between these two methods were considered as evidence of a causal effect. The Bonferroni corrected p-value<0.05 based on the number of all MR tests performed was used to determine significance. The MR analysis was conducted using the R package “ivmodel”[22].

## 3. Results

### 3.1 Participant characteristics

Characteristics of the WRAP and Wisconsin ADRC participants can be found in Table 1. Among 161 WRAP participants, the mean baseline age and education level were 62.1 and 16.2 years, respectively. The mean age and years of education in the Wisconsin ADRC were 58.1 and 16.2, respectively. Females comprised 65.2% of WRAP participants and 68.8% of the Wisconsin ADRC. The mean values of each biomarker are also listed in Table 1.

**Table 1.** Sample characteristics of WRAP and Wisconsin ADRC participants.

|  | **WRAP\***<br>**N=161** |  | **Wisconsin**<br>**ADRC**<br>**N=154** |  |
| --- | --- | --- | --- | --- |
|  | **Mean** | **SD** | **Mean** | **SD** |
| **Age** | 62.1 | 6.5 | 58.1 | 5.5 |
| **Years of education** | 16.2 | 2.2 | 16.2 | 2.3 |
| **P-tau** | 17.5 | 6.4 | 15.9 | 5.8 |
| **T-tau** | 200.6 | 67.3 | 184.5 | 69.3 |
| **Aβ42** | 895.4 | 369.4 | 942.6 | 363.1 |
| **Aβ40** | 14336.1 | 4497.9 | 13897.4 | 4742.3 |
| **NfL** | 87.2 | 38.0 | 83.3 | 80.7 |
| **Neurogranin** | 798.6 | 307.5 | 728.2 | 286.7 |
| **YKL-40** | 144.6 | 48.3 | 128.7 | 39.1 |
| **S100b** | 1.1 | 0.3 | 1.2 | 0.3 |
| **GFAP** | 8.6 | 2.9 | 8.6 | 3.3 |
| **sTREM2** | 7.9 | 2.4 | 7.6 | 2.1 |
| **IL-6** | 4.3 | 2.6 | 5.2 | 3.7 |
| **α-synuclein** | 157.4 | 65.1 | 146.2 | 64.1 |
|  | **N** | **%** | **N** | **%** |
| **Female** | 105 | 65.2 | 106 | 68.8 |
| **Male** | 56 | 34.8 | 48 | 31.2 |

\* The summary statistics of WRAP were based on the baseline measures.

### 3.2 MWAS

The significant MWAS results in WRAP and the Wisconsin ADRC are summarized in Figure 1. In WRAP, a large number of CSF metabolites reached the significance threshold after FDR correction [Figure 1. (a)]. 47 metabolites were associated with P-tau, 56 were associated with T-tau, 58 were associated with Aβ42, 80 were associated with Aβ40, 65 were associated with NfL, and 62 were associated with neurogranin. However, no metabolites were associated with the ratio of Aβ42/40 or IL-6. Many of the metabolites that were significant in WRAP were also significant in the Wisconsin ADRC [Figure 1. (b)]. For example, among 47 significant metabolites for P-tau in WRAP, 40 metabolites were also significant in the Wisconsin ADRC. Table 2 shows the replication results for the top 10 significant CSF metabolite-biomarker associations (if there were 10 or more significant metabolites) in the Wisconsin ADRC. For example, the top three metabolites associated with P-tau and T-tau were 1-palmitoyl-2-stearoyl-GPC (16:0/18:0), N-acetylneuraminate, and C-glycosyltryptophan. N-acetylneuraminate and 1,2-dipalmitoyl-GPC (16:0/16:0) were the top two metabolites associated with Aβ42 and Aβ40. The top three metabolites associated with NfL were N-acetylthreonine, N-acetylalanine, and beta-citrylglutamate. N-acetylneuraminate, C-glycosyltryptophan, and N6-succinyladenosine were the top three metabolites for neurogranin. N-acetylneuraminate, 1,2-dipalmitoyl-GPC (16:0/16:0), and stearoyl sphingomyelin (d18:1/18:0) were the top three metabolites for YKL40. Stearoyl sphingomyelin (d18:1/18:0), 1-stearoyl-2-docosahexaenoyl-GPC (18:0/22:6), and 1-palmitoyl-2-oleoyl-GPC (16:0/18:1) were the top three metabolites associated with S100b. Only six metabolites were associated with GFAP, and the top three were 1,2-dipalmitoyl-GPC (16:0/16:0), 1-palmitoyl-2-stearoyl-GPC (16:0/18:0), and beta-citrylglutamate. For sTREM2, the top metabolites were stearoyl sphingomyelin (d18:1/18:0), 1,2-dipalmitoyl-GPC (16:0/16:0), and palmitoyl sphingomyelin (d18:1/16:0). Finally, for α-synuclein, the top three metabolites were 1-palmitoyl-2-stearoyl-GPC (16:0/18:0), 1,2-dipalmitoyl-GPC (16:0/16:0), and N-acetylneuraminate. The full results of WRAP and the Wisconsin ADRC can be found in Supplemental Tables 2-25. The association patterns between significant CSF metabolites and NTK biomarkers are provided in the Figure 2. The summary of the number of significant associations and the name of NTK biomarkers that were replicated in the Wisconsin ADRC are presented in Supplemental Table 26. Most of the significant metabolites were lipids, amino acids, and carbohydrates. For example, the lipid, 1,2-dipalmitoyl-GPC (16:0/16:0), the amino acid, beta-citrylglutamate, and the carbohydrate N-acetylneuraminate were strongly associated with almost every CSF NTK biomarker of AD. On the contrary, amino acids like kynurenate and proline were only significantly associated with α-synuclein.

**Figure 1.**
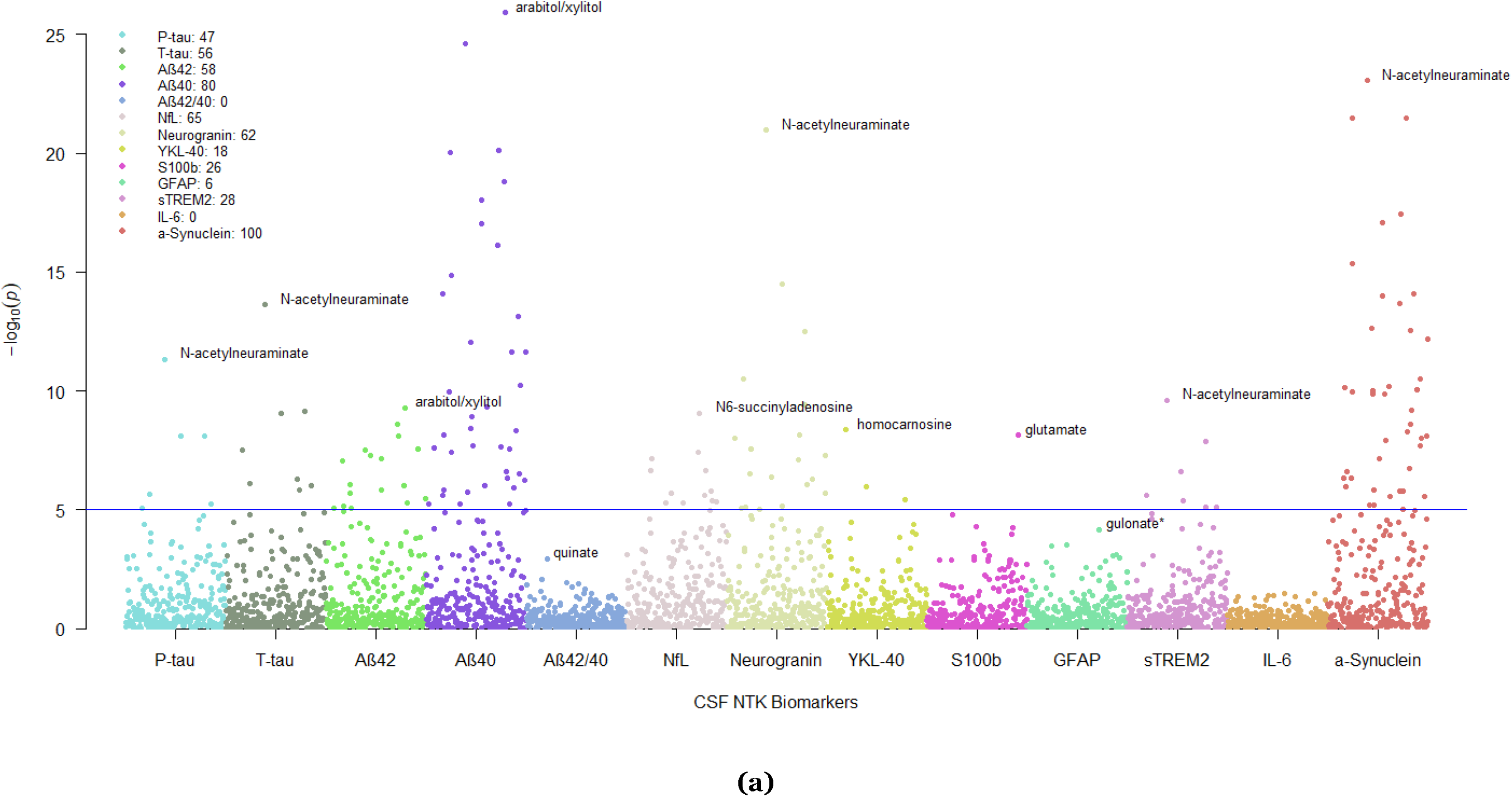
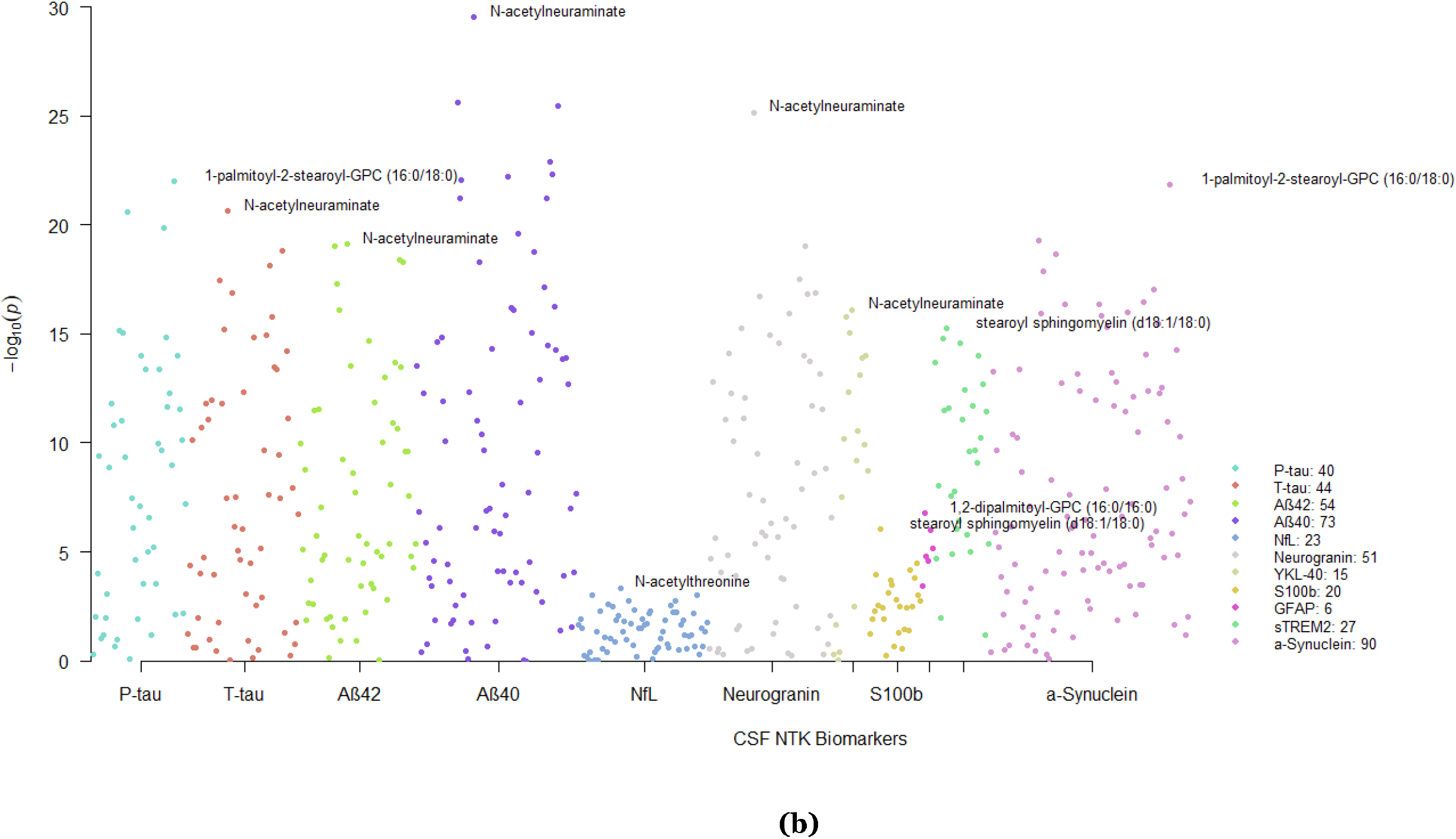
Manhattan plots of MWAS results between CSF metabolites and CSF NTK biomarkers. Each dot represents a metabolite and the different colors represent the CSF NTK biomarkers (x-axis) in (a) WRAP and (b) the Wisconsin ADRC (only significant metabolites after FDR correction in WRAP were included). The -log10(p-value) is shown on the y-axis. The legend box indicates the number of metabolites that were significant after FDR correction for each NTK biomarker.

**Figure 2.**
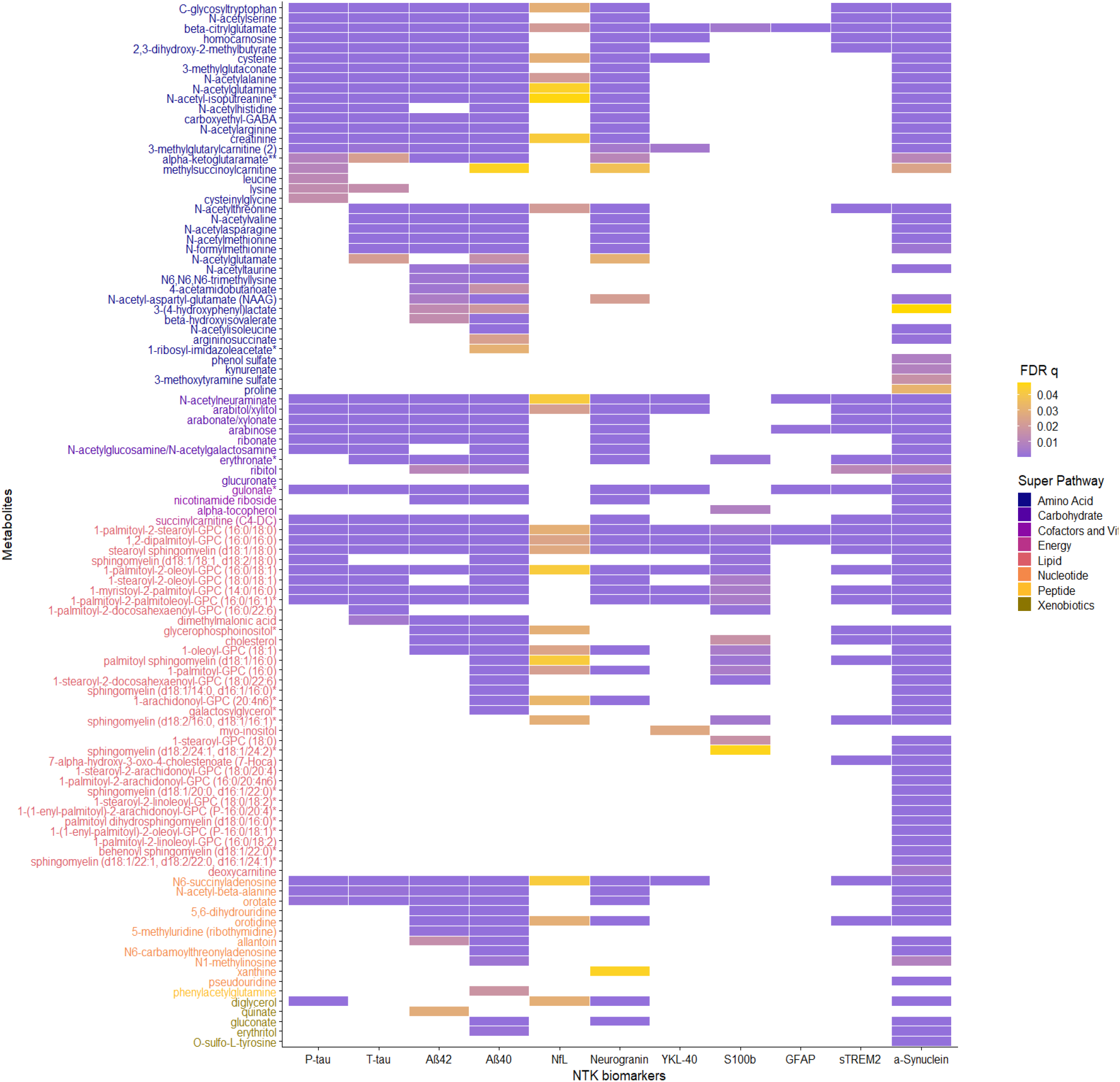
The association patterns between significant CSF metabolites and CSF NTK biomarkers in Wisconsin-ADRC. Each cell represents the association of a CSF metabolites with a biomarker. The color scale indicates the magnitude of the FDR q values. The metabolites are also grouped and colored based on their super pathway.

**Table 2.**
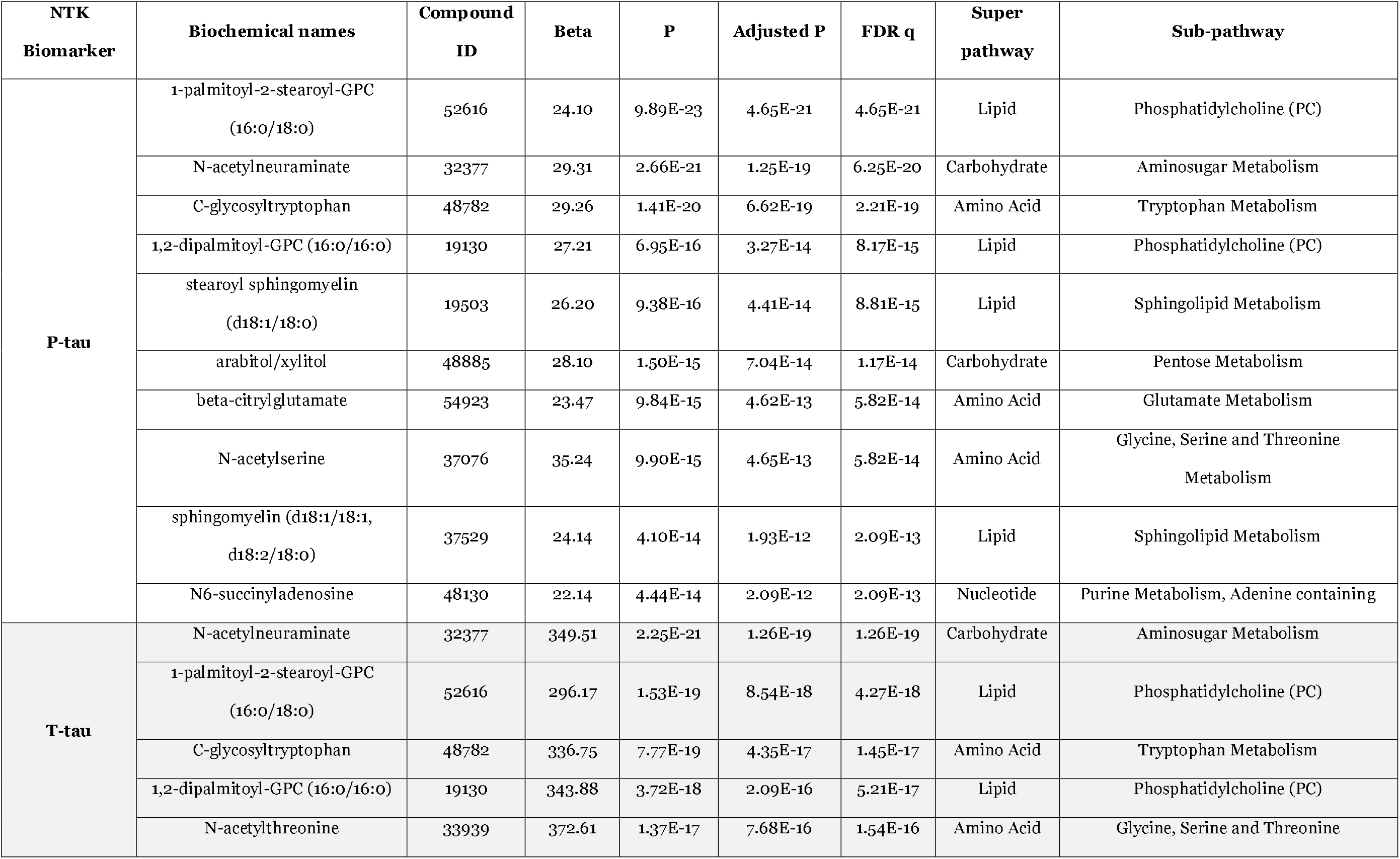
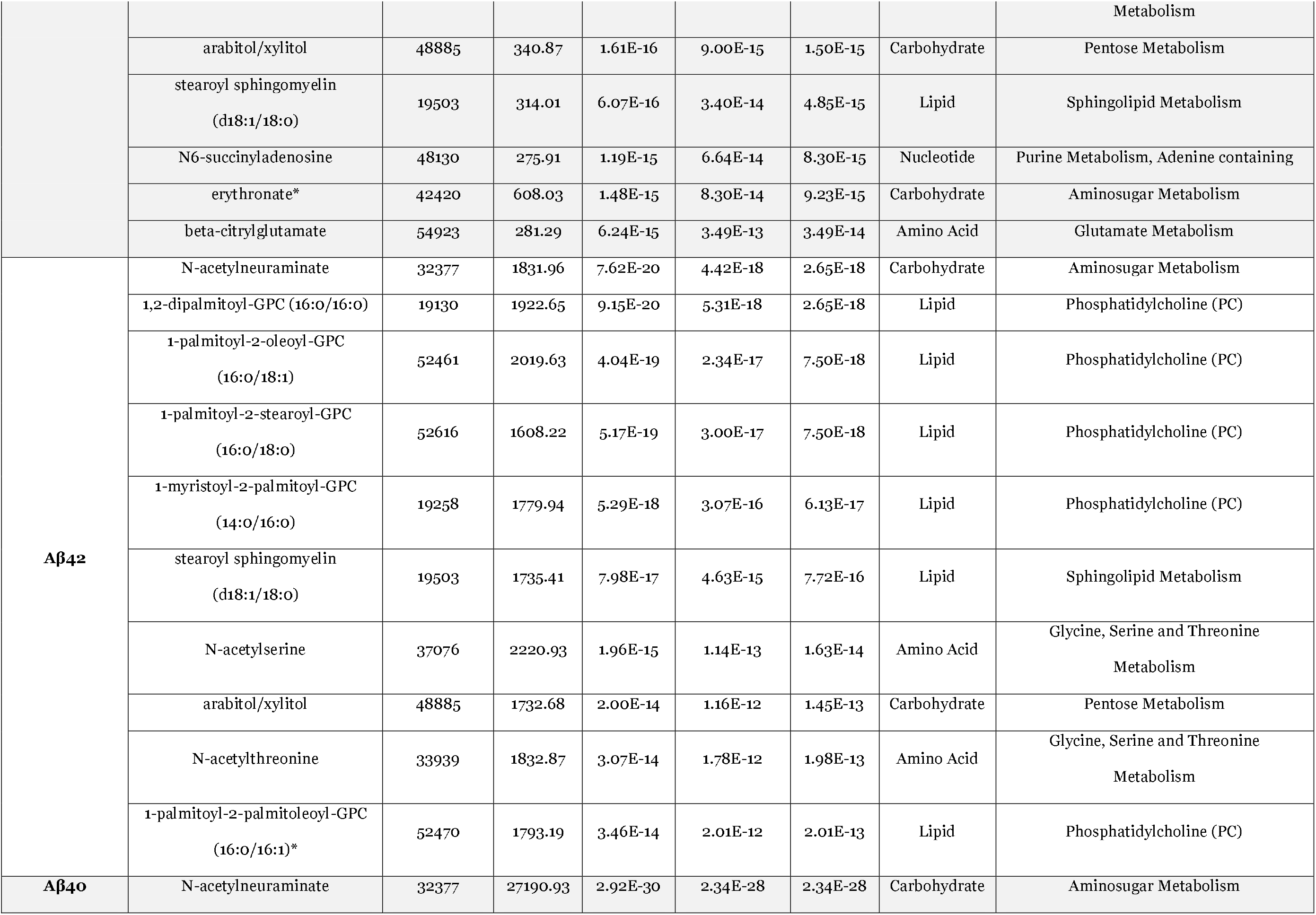
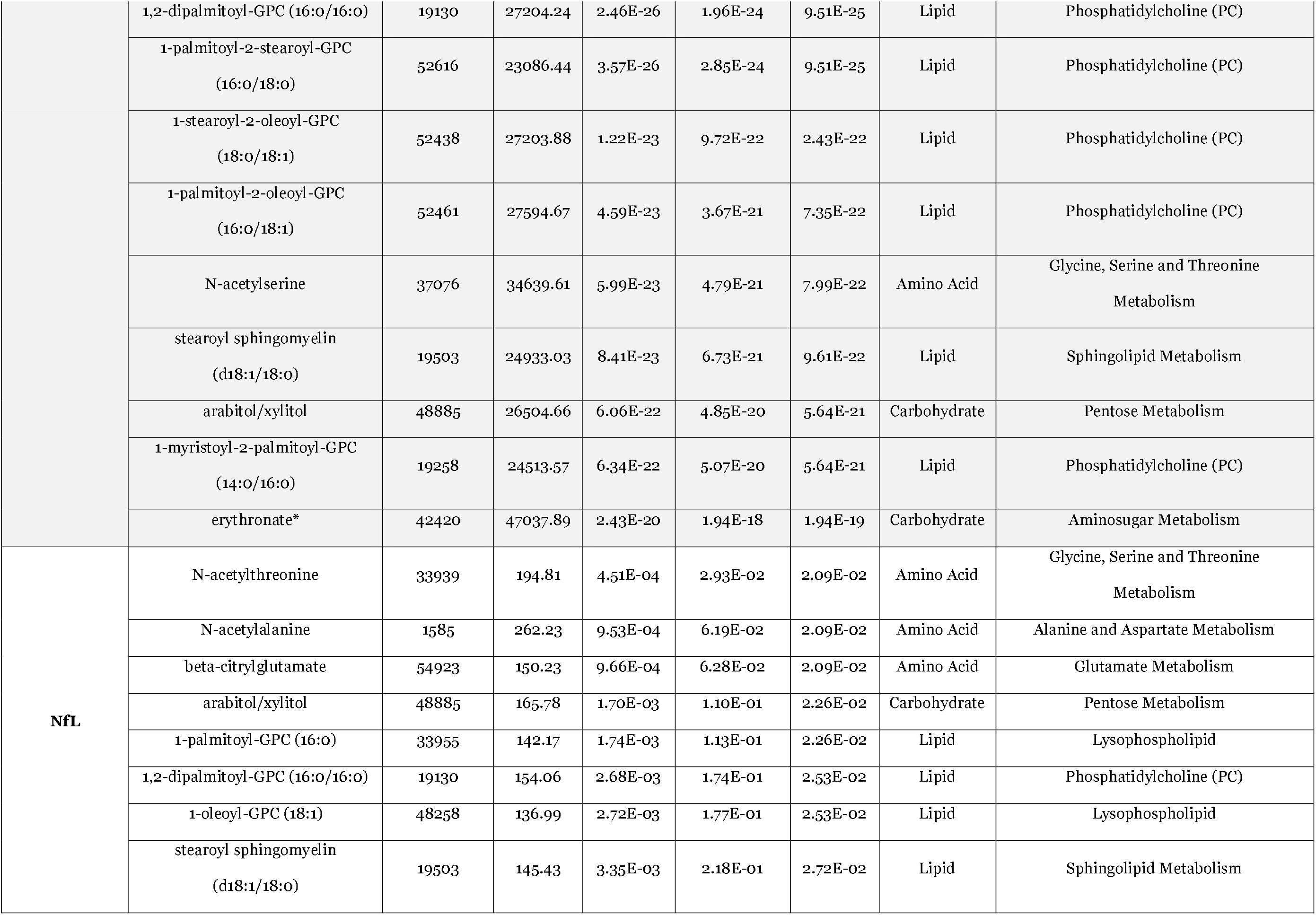
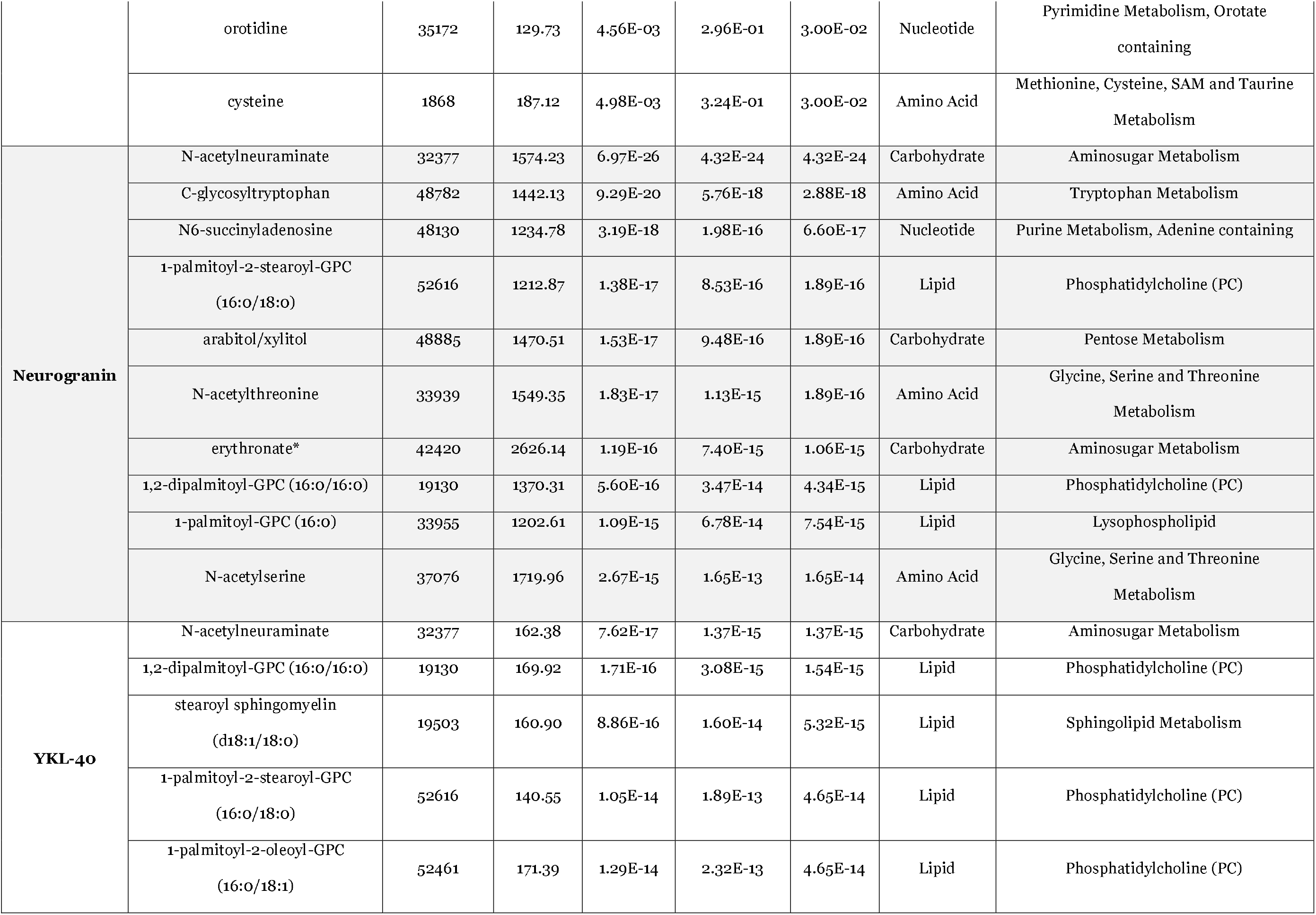
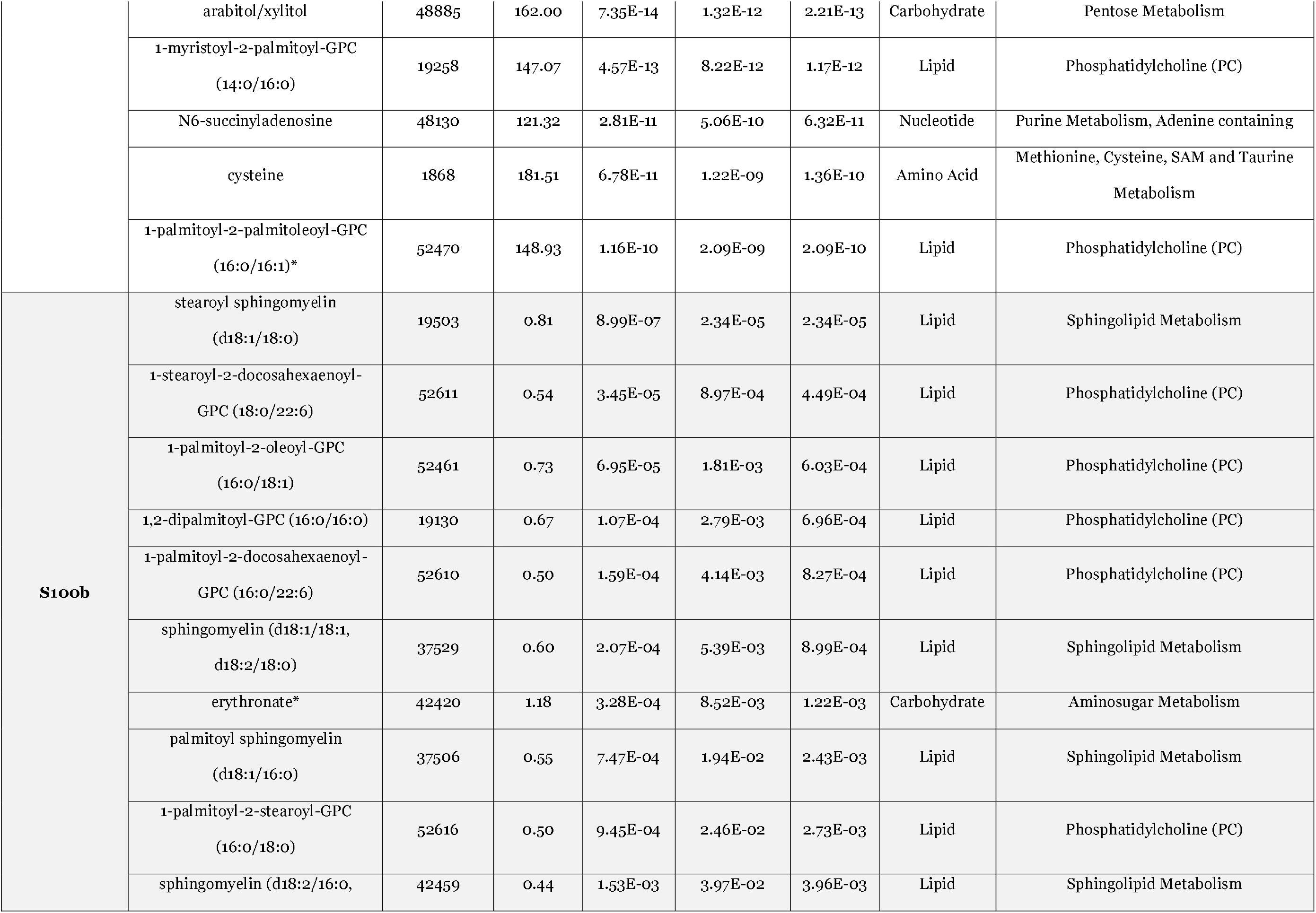
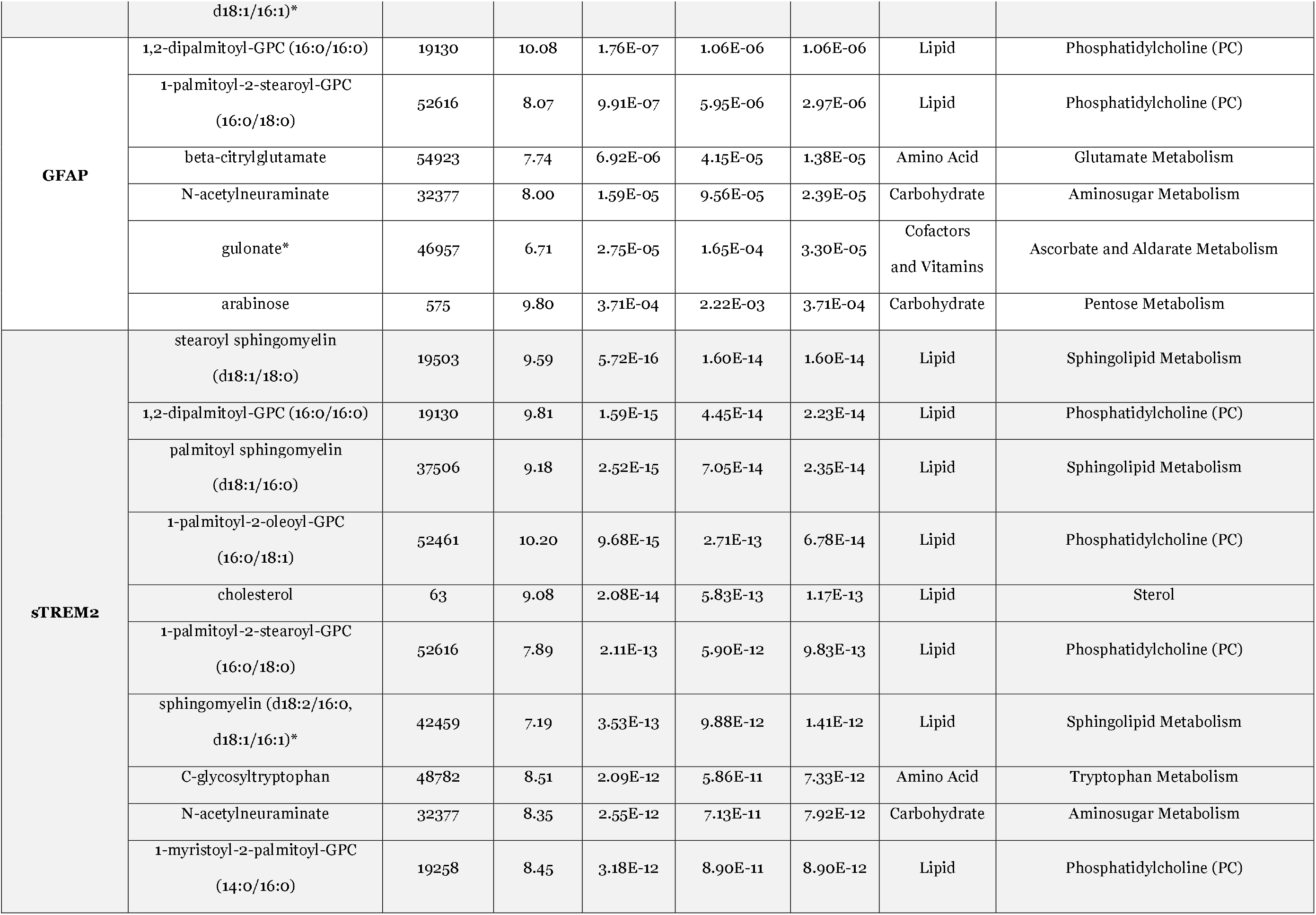
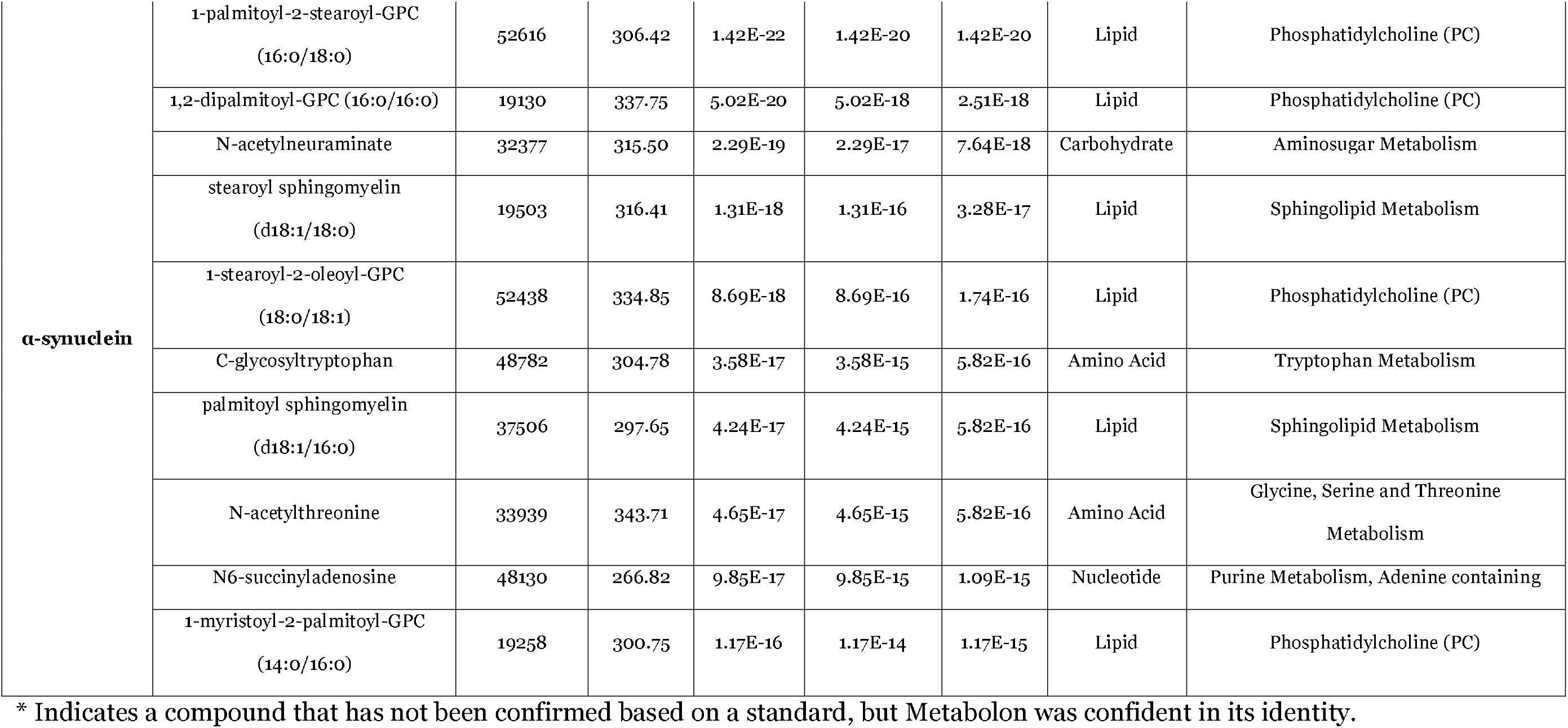
Top 10 significant CSF metabolites associated with each NTK biomarker in WRAP and replicated in the Wisconsin ADRC.

The functional pathways for replicated significant metabolites with known human metabolome database (HMDB) IDs for each CSF NTK biomarker are shown in Supplemental Table 27. Two significant metabolites, 1,2-dipalmitoyl-GPC (16:0/16:0) and 1-oleoyl-GPC (18:1), were enriched in the glycerophospholipid metabolism pathway for most biomarkers. Other pathways such as pyrimidine metabolism (including orotate and orotidine), ascorbate and aldarate metabolism (including gulonate and glucuronate), arginine biosynthesis (including N-acetylglutamate and argininosuccinate), and pentose and glucuronate interconversions (also including gulonate and glucuronate) may also be of interest.

### 3.3 Prediction performance and elastic net regression results

The prediction performance of replicated significant metabolites was measured by r^2^ and presented in Table 3. The r^2^ of the base models, which only included the demographic variables, ranged from 0.01 to 0.25. Adding the replicated significant metabolites increased the r^2^ substantially for each biomarker, ranging from 0.13 to 0.94. The elastic net regression further prioritized candidate metabolites associated with each biomarker. For example, 22 of the original 40 significant metabolites were selected by the elastic net as important independent metabolites for P-tau. Initially, 40 significant metabolites explained about 72% of the variance in P-tau; the 22 elastic net-selected metabolites still explained 70% of the variance.

**Table 3.** Prediction performance (r^2^) of metabolites in the combined cohort of WRAP and Wisconsin ADRC.

| **NTK biomarkers** | **Base model $r^2$ (sample size\*)** | **Number of input metabolites** | **Metabolite model $r^2$ (sample size\*)** | **Number of elastic net-selected metabolites** | **Elastic net-selected metabolites model $r^2$ (sample size\*)** |
| --- | --- | --- | --- | --- | --- |
| **P-tau** | 0.11 (n=295) | 40 | 0.72 (n=226) | 22 | 0.70 (n=232) |
| **T-tau** | 0.10 (n=296) | 44 | 0.71 (n=236) | 17 | 0.69 (n=281) |
| **Aβ42** | 0.01 (n=296) | 54 | 0.55 (n=220) | 43 | 0.54 (n=222) |
| **Aβ40** | 0.04 (n=293) | 73 | 0.85 (n=162) | 70 | 0.83 (n=166) |
| **NfL** | 0.10 (n=297) | 23 | 0.23(n=280) | 13 | 0.22 (n=290) |
| **Neurogranin** | 0.07 (n=297) | 51 | 0.78 (n=245) | 32 | 0.76 (n=267) |
| **YKL-40** | 0.25 (n=297) | 15 | 0.57 (n=281) | 10 | 0.56 (n=288) |
| **S100** | 0.05 (n=296) | 20 | 0.22 (n=281) | 4 | 0.19 (n=293) |
| **GFAP** | 0.17 (n=297) | 6 | 0.35 (n=282) | 5 | 0.35(n=288) |
| **sTREM2** | 0.05 (n=297) | 27 | 0.52 (n=276) | 14 | 0.50 (n=283) |
| **α-synuclein** | 0.03 (n=297) | 90 | 0.97 (n=105) | 27 | 0.68 (n=218) |

Variables included in the base model were age, sex, years of education and cohort.
Variables included in the metabolite model were age, sex, years of education, cohort and all replicated significant metabolites for each biomarker.
Variables included in the metabolite model were age, sex, years of education, cohort and elastic net-selected metabolites.
\* The sample sizes were different because of the missingness of metabolites.

### 3.4 Mendelian randomization

According to the F statistics, we employed the LIML method for MR. The full results of the test statistics are provided in Supplemental Table 28. After checking for consistency of the CIs for the LIML and CLR methods, the significant and consistent MR results are displayed in Table 4, showing metabolites with a potential causal effect on the NTK biomarker based on instrumental variables formed by genome-wide significant SNPs. For example, we observed a positive causal association between palmitoyl sphingomyelin (d18:1/16:0) and sTREM2.

**Table 4.**
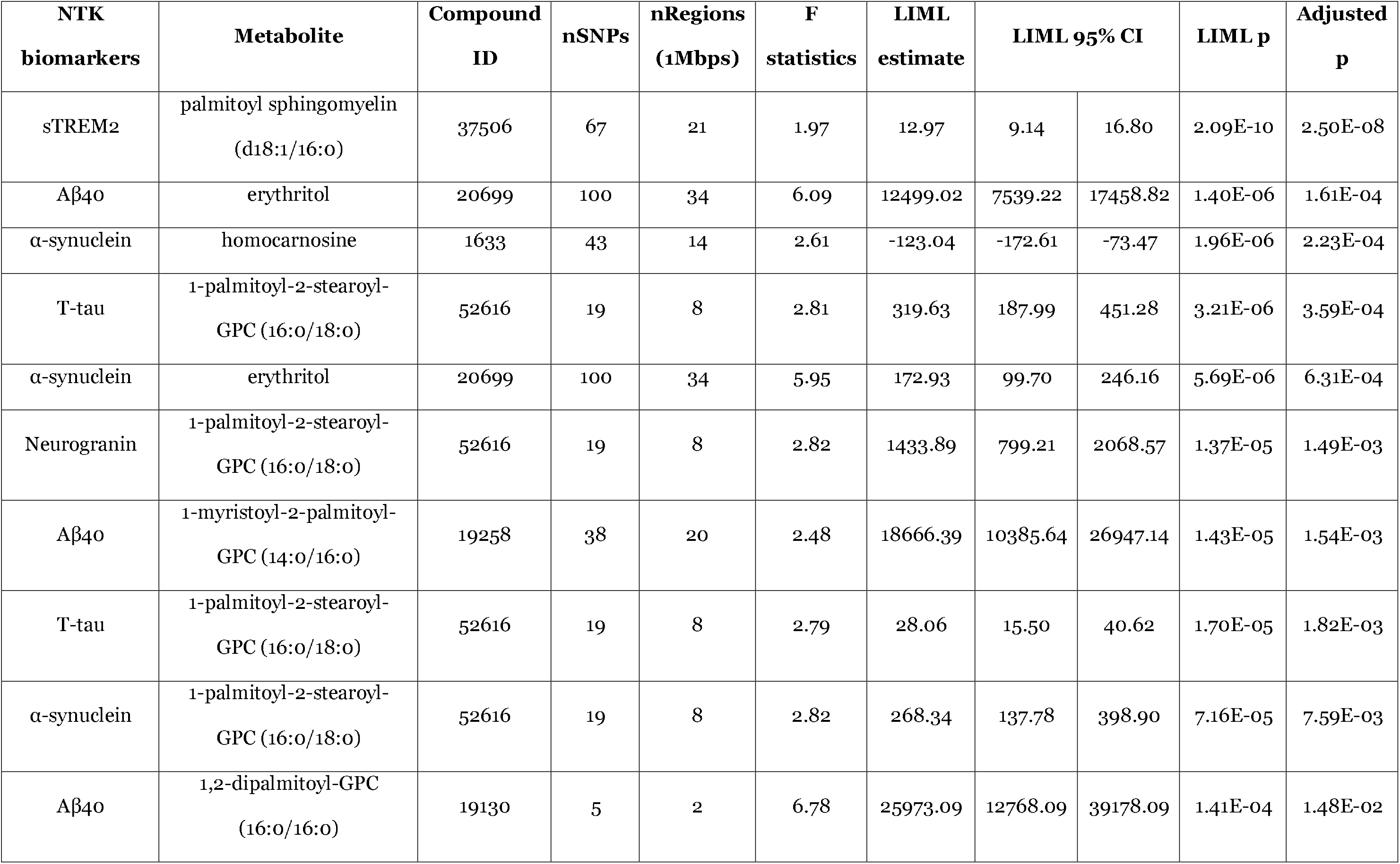
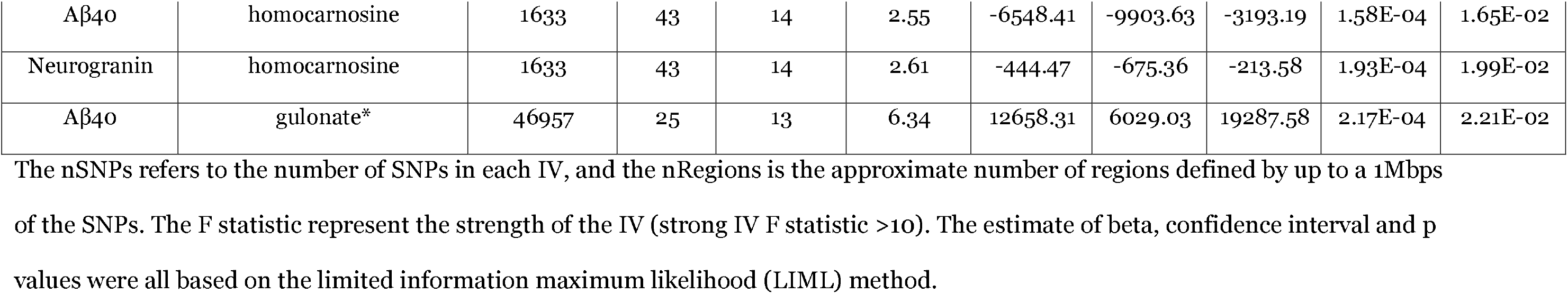
Significant Mendelian randomization results after Bonferroni correction.

## 4. Discussion

In this analysis, we tested the associations between CSF metabolites and CSF NTK biomarkers representing different pathologies of AD in initially cognitively-unimpaired individuals. Significant metabolites were identified in the WRAP cohort using linear mixed effects regression and most of the metabolites were replicated in the Wisconsin ADRC cohort. The elastic net regression method reduced the number of CSF metabolites by selecting the important and independent metabolites for each CSF biomarker. This provides a smaller, more practical set of metabolites to focus on in future research. The results of the MR analyses suggested several metabolites that may play a causal role in AD pathology. A detailed look into these associations, such as the contributing genes and their corresponding functions, is worth exploring.

We have identified and replicated multiple CSF metabolites that were associated with CSF NTK biomarkers for AD pathology; most of these CSF metabolites were lipids, particularly sphingolipids, phosphatidylcholines, and lysophospholipids, which are all types of phospholipids. Phospholipids are a class of lipids that construct the cellular membranes and are involved in many complex activities of membrane proteins, receptors, enzymes, and ion channels in the cell or at the cell surface[23]. In the neurodegenerative brain, e.g., in the AD brain, which has suffered extensive damage, the compromise of the membrane functions is expected, explaining how phospholipids may be involved in AD pathology[24]. Previous studies have demonstrated that various phospholipids such as phosphatidylcholines, sphingolipids, glycerophospholipids, and lysophospholipids have changed in the AD patient’s brain, CSF and blood when compared to healthy controls[23,25,26]. For example, a serum metabolomics study conducted by González-Domínguez *et al.*[25] showed that the concentration of numerous phosphatydyl lipids, like 1,2-dipalmitoyl-GPC (16:0/16:0), 1-palmitoyl-2-linoleoyl-GPC (16:0/18:2), and 1-palmitoyl-2-oleoyl-GPC (16:0/18:1), and lysophosphatidylcholines, like 1-palmitoyl-GPC (16:0) and 1-stearoyl-GPC (18:0), were different in AD versus healthy controls. The 1,2-dipalmitoyl-GPC (16:0/16:0) phosphatidylcholine has also been suggested as one of three serum metabolites to predict AD development in MCI individuals[27]. Another brain metabolomics study found that higher levels of palmitoyl sphingomyelin (d18:1/16:0) and sphingomyelin (d18:1/18:1, d18:2/18:0) were associated with the severity of AD pathology at autopsy and AD progression across prodromal and preclinical stages[28]. The stearoyl sphingomyelin (d18:1/18:0) was also significantly changed in the CSF with “AD-like pathology” that was dichotomized by Aβ42, T-tau, and P-tau levels[6]. In summary, our results confirmed the importance of the previously identified lipids but also provided novel lipid findings for AD pathologies beyond the major established ones.

Another class of metabolites that are of potential interest are several carbohydrates like N-acetylneuraminate, arabitol/xylitol, arabinose, and erythronate. Among them, N-acetyleneuraminate, also known as sialic acid, had a significant effect on most NTK biomarkers. In addition to our study, a previous study conducted by Nagata *et al.*[29] in 2018 also showed that CSF N-acetylneuraminate was significantly increased in AD when compared to patients with idiopathic normal pressure hydrocephalus and was positively correlated with CSF P-tau (r=0.55), as it was in our study. N-acetyleneuraminate is an acetyl derivative of the amino sugar neuraminic acid, which occurs in many glycoproteins, glycolipids, and polysaccharides. Specifically, it is a functional and structural component of gangliosides, which are found predominantly in the nervous system and are abundant in the brain, especially in the grey matter[30]. Studies have shown that gangliosides play important roles in AD. For example, it has been suggested that GM1-ganglioside binds to Aß, and the resulted GAß has the capability to accelerate Aß assembly[31] and is the endogenous seed for amyloid fibral in the AD brain[32]. The gangliosides also have important roles in organizing the lipid rafts, which integrate numerous types of lipid proteins involved in cell signaling, cell-cell adhesion, and intracellular vesicular trafficking[29] and contain many AD-associated proteins such as amyloid precursor protein (APP)[33]. Furthermore, the gene *CD33*, which belongs to the sialic-acid-binding immunoglobulin-like lectin family, has been reported as a strong genetic locus associated with AD by GWASs[34–36] and has been suggested to impair the microglia-mediated Aβ clearance[37–39]. Erythronate (erythronic acid) was previously identified as the main hallmark of pentose–phosphate pathway defects[40], and consistent with abnormal function of pentose–phosphate pathway in certain regions of the AD-brain[41], and the upregulation of the pentose–phosphate pathway was reported in a previous study of mild cognitive impairment (MCI) participants that later progressed to AD[42].

As mentioned above, a couple of metabolites were common to most of the AD pathologies defined by the CSF NTK biomarkers. On the contrary, some metabolites were unique to specific NTK biomarkers. For example, lipids like 1-palmitoyl-2-linoleoyl-GPC (16:0/18:2), 1-stearoyl-2-arachidonoyl-GPC (18:0/20:4), sphingomyelin (d18:1/20:0, d16:1/22:0) and sphingomyelin (d18:1/22:1, d18:2/22:0, d16:1/24:1) were only associated with α-synuclein. These metabolites may be helpful to study synaptic dysfunction and could potentially be used as biomarkers to differentiate AD pathologies.

By utilizing Mendelian randomization, we found causal evidence for several of the associations between CSF metabolites and CSF NTK biomarkers. Among these metabolites, most of them were lipids, with some amino acids and cofactors/vitamins, and a xenobiotic metabolite, erythritol. Another metabolite of interest, homocarnosine, is an inhibitory neuromodulator synthesized in the neuron from gamma-aminobutyric acid (GABA) and histidine[43]. The level of human CSF homocarnosine declines drastically with age [44] and was suggested to be related to AD through CSF protein glycation[45]. At the same time, GABA also plays an important role in the brain and may be related to AD[46].

This study has some limitations. First, the analysis only included non-Hispanic white individuals, so the results may not extrapolate to other racial/ethnic groups. Second, the sample sizes of both the WRAP and Wisconsin ADRC cohorts were relatively small and will need to be replicated in a larger independent sample. The significant associations between a number of metabolites and both Aβ42 and Aβ40, but not with Aβ42/40 may indicate that the metabolites associated with Aβ42 and Aβ40 only influence the production of amyloid in general versus clearance of the pathological form, Aβ42. In general, the research confirmed that several novel metabolites changed along with AD CSF biomarkers and extended several developing and understudied AD pathologies, e.g., synaptic dysfunction, based on untargeted CSF metabolomics and will expand our knowledge of the biological mechanisms behind AD.

## Data Availability

The data underlying this article can be requested through the WRAP and Wisconsin ADRC Application for any qualified investigator.

## Acknowledgments

The authors especially thank the WRAP and Wisconsin ADRC participants and staff for their contributions to the studies. Without their efforts, this research would not be possible. This study was supported by the National Institutes of Health (NIH) grants [R01AG27161 (Wisconsin Registry for Alzheimer Prevention: Biomarkers of Preclinical AD), R01AG054047 (Genomic and Metabolomic Data Integration in a Longitudinal Cohort at Risk for Alzheimer’s Disease), R01AG037639 (White Matter Degeneration: Biomarkers in Preclinical Alzheimer’s Disease), R21AG067092 (Identifying Metabolomic Risk Factors in Plasma and Cerebrospinal Fluid for Alzheimer’s Disease), and P30AG062715 (Wisconsin Alzheimer’s Disease Research Center Grant)], the Helen Bader Foundation, Northwestern Mutual Foundation, Extendicare Foundation, State of Wisconsin, the Clinical and Translational Science Award (CTSA) program through the NIH National Center for Advancing Translational Sciences (NCATS) grant [UL1TR000427], and the University of Wisconsin-Madison Office of the Vice Chancellor for Research and Graduate Education with funding from the Wisconsin Alumni Research Foundation. Computational resources were supported by core grants to the Center for Demography and Ecology [P2CHD047873] and the Center for Demography of Health and Aging [P30AG017266].

HZ is a Wallenberg Scholar supported by grants from the Swedish Research Council (#2018-02532), the European Research Council (#681712), Swedish State Support for Clinical Research (#ALFGBG-720931), the Alzheimer Drug Discovery Foundation (ADDF), USA (#201809-2016862), the AD Strategic Fund and the Alzheimer’s Association (#ADSF-21-831376-C, #ADSF-21-831381-C and #ADSF-21-831377-C), the Olav Thon Foundation, the Erling-Persson Family Foundation, Stiftelsen för Gamla Tjänarinnor, Hjärnfonden, Sweden (#FO2019-0228), the European Union’s Horizon 2020 research and innovation programme under the Marie Skłodowska-Curie grant agreement No 860197 (MIRIADE), European Union Joint Program for Neurodegenerative Disorders (JPND2021-00694), and the UK Dementia Research Institute at UCL.

KB is supported by the Swedish Research Council (#2017-00915), ADDF, USA [#RDAPB-201809-2016615], the Swedish Alzheimer Foundation [#AF-742881], Hjärnfonden, Sweden [#FO2017-0243], the Swedish state under the agreement between the Swedish government and the County Councils, the ALF-agreement [#ALFGBG-715986], and European Union Joint Program for Neurodegenerative Disorders [JPND2019-466-236], and the Alzheimer’s Association 2021 Zenith Award (ZEN-21-848495).

We thank the University of Wisconsin Madison Biotechnology Center Gene Expression Center for providing Illumina Infinium genotyping services and Roche for providing the NTK kits for this study. COBAS, COBAS E and ELECSYS are trademarks of Roche.

